# Context-Dependent Age-Group performance hierarchies limit fairness interventions in PPG-based heart rate prediction

**DOI:** 10.64898/2026.06.04.26352929

**Authors:** Lovely Yeswanth Panchumarthi, Saurabh Kataria, Yi Wu, Xiao Hu, Alex Fedorov, Hyunjung Gloria Kwak

## Abstract

**Background:** Fairness-aware machine learning increasingly targets demographic performance disparities in clinical prediction, yet whether standard bias mitigation strategies genuinely improve equity in physiological signal analysis remains unclear. Age-based disparities in photoplethysmography (PPG)-based heart rate prediction present a particular challenge, as age-related performance differences may reflect context-dependent physiological structure rather than correctable artifacts.

**Methods:** We evaluated three fairness interventions — inverse-frequency weighting (IF), Group Distributionally Robust Optimization (GroupDRO), and adversarial debiasing (ADV) — applied via fine-tuning of a PPG foundation model across three clinical datasets spanning intensive care unit, laboratory, and consumer wearable contexts. Outcomes were assessed using a 2 *×* 2 framework classifying each intervention–dataset combination by the joint direction of change in mean absolute error (MAE) and fairness gap (FG) across age groups, yielding four outcome types: genuine improvement (G), leveling down (L), selective benefit (S), and both worse (W).

**Results:** Across nine intra-domain conditions, no intervention simultaneously improved both MAE and FG (0/9 genuine improvement). The dominant pattern was leveling down (5/9): FG decreased but was accompanied by MAE degradation, indicating that apparent fairness gains were achieved at the cost of overall predictive performance. Age-group difficulty ordering varied across clinical contexts at baseline and was not preserved under intervention.

In 18 cross-domain transfer conditions, genuine improvement was rare (4/18) and observed exclusively in non-MIMIC source configurations; models fine-tuned on MIMIC-sourced data yielded no genuine improvements (0/6). Embedding-level representation changes following fine-tuning did not reliably predict fairness outcomes.

**Conclusions:** Age-based fairness interventions in PPG heart rate prediction indicate a leveling-down pattern rather than genuine equity improvement, suggesting that age-related performance gaps reflect context-dependent physiological structure not fully addressable through standard bias mitigation. Cross-domain transfer further amplifies this instability. These findings suggest that fairness evaluation frameworks for age-stratified physiological prediction should account for context-dependent performance structure rather than treating observed gaps as correctable bias.

**Author summary:** When machine learning models predict health outcomes less accurately for some demographic groups than others, a common response is to apply “fairness” corrections during model training. These methods assume that performance gaps arise from fixable problems in the data or algorithm. We asked whether this assumption holds for

age-based disparities in heart rate prediction from wrist-worn and clinical sensors. Across three clinical settings — consumer wearables, laboratory recordings, and intensive care — we found that the same age group could be the easiest or hardest to predict depending on the setting, reflecting genuine physiological differences rather than a fixable flaw. When we applied three standard fairness methods, none simultaneously improved accuracy and fairness; instead, most appeared to improve fairness only by making predictions worse for everyone. Models corrected in one setting also performed poorly when moved to another. These results suggest that not all demographic performance gaps are correctable, and that reporting fairness improvements without checking whether overall quality declined can be misleading.

## Introduction

Photoplethysmography (PPG)-based heart rate monitoring spans intensive care units, laboratory assessments, and consumer wearable devices, creating deployment contexts with fundamentally different physiological characteristics [1]. As pretrained foundation models for PPG analysis — such as GPT-PPG [2], SiamQuality [3], and PaPaGei [4] — become available for downstream adaptation through fine-tuning [5, 6], the question of whether fine-tuned models perform equitably across demographic subgroups has gained urgency, particularly as pretrained demographic associations can persist through or be amplified by fine-tuning [7, 8].

The standard approach to demographic performance disparities in clinical machine learning applies fairness interventions — demographic reweighting, distributionally robust optimization [9], or adversarial debiasing [10] — during training to equalize subgroup performance. These methods share an implicit assumption: that demographic performance gaps represent correctable artifacts arising from data imbalance, representational bias, or learnable shortcuts [11, 12, 13]. Under this assumption, the appropriate remedy involves rebalancing the contribution of underperforming groups during optimization.

However, a growing body of evidence suggests that fairness interventions in clinical ML frequently reduce inter-group disparities by degrading performance for all groups — a phenomenon formalized as “leveling down” [14]. Zhang et al. [15] found that methods achieving group fairness on chest X-ray classifiers did so by worsening predictions across subgroups, regardless of the specific fairness algorithm applied. Pfohl et al. [16] characterized this as “nearly-universal degradation” of within-group metrics under fairness constraints. Yang et al. [17] further demonstrated that fairness gains achieved within one data distribution do not transfer to new deployment settings, characterizing within-distribution corrections as “locally optimal” but not globally so. These findings raise a deeper question: whether the assumption that performance gaps represent correctable artifacts holds uniformly across demographic attributes and clinical contexts.

Age may be a demographic axis for which this assumption is particularly problematic. Unlike measurement-linked attributes such as skin tone — where performance gaps have been linked to device calibration artifacts [18, 19] — age interacts with physiological prediction difficulty in context-dependent ways. In healthy populations, heart rate variability declines with age as autonomic regulation narrows [20, 21], potentially making elderly individuals easier prediction targets at rest. In critically ill populations, multiple organ dysfunction can overwhelm normal age-dependent autonomic patterns [22], and hemodynamic instability increases with age [23], making elderly patients the most difficult prediction targets. The same age group may thus shift from lowest to highest prediction error depending on the clinical context — a structural property that fairness interventions designed to correct artifacts are not equipped to address.

Despite growing recognition of these challenges, prior fairness evaluations have predominantly operated within single-domain settings where the artifact assumption plausibly holds [18, 24, 25]. Benchmarking efforts such as MEDFAIR [26] have shown that bias mitigation algorithms often yield mixed results even in settings where algorithmic bias is presumed, and cross-site studies [17] have documented the fragility of fairness gains under distribution shift. Liu et al. [27] have further argued that performance differences across demographic groups may reflect clinically meaningful variation rather than correctable bias, cautioning against treating all observed disparities as targets for algorithmic mitigation. Yet to our knowledge, prior work has not examined how fairness interventions behave when the performance hierarchy they seek to correct is itself context-dependent.

We evaluate three established fairness interventions — inverse-frequency weighting (IF), Group Distributionally Robust Optimization (GroupDRO), and adversarial debiasing (ADV) — applied during fine-tuning of a pretrained PPG model for heart rate prediction across three datasets: consumer wearables (DaLiA), laboratory recordings (BUT-PPG), and intensive care monitoring (MIMIC). We assess nine within-domain and eighteen cross-domain method–context combinations, decomposing outcomes by joint direction of accuracy and fairness change to distinguish genuine improvement from leveling down. Our results indicate that age-group performance hierarchies reverse across clinical contexts, that fairness gap reduction within domains occurs predominantly through accuracy degradation for lower-error groups, and that cross-domain fairness transfer depends on alignment between source and target physiological structures rather than on the choice of intervention.

## Methods

### Pretrained Model and Fine-Tuning Protocol

We adopted the experimental protocol established in our companion study [28], which introduced a bias-aware fine-tuning framework for GPT-PPG [2], a decoder-only transformer pretrained on over 200 million PPG waveform segments from critical care settings. Full architectural details, pretraining procedures, and fine-tuning hyperparameters are described in our companion study and Chen et al. [2, 28]. Briefly, we fine-tuned the 19M-parameter GPT-PPG configuration for heart rate prediction as a regression task using L1 loss with Logit-Laplace auxiliary regularization, Adam optimization with cosine annealing, and full-layer updates for up to 100 epochs. All PPG signals were resampled to 40 Hz and segmented into one-second non-overlapping windows.

### Datasets and Age Stratification

We used three publicly available PPG datasets spanning distinct acquisition contexts: DaLiA [29], containing wrist-worn wearable recordings at 64 Hz during daily activities including walking, cycling, and desk work; BUT-PPG [30], containing 10-second smartphone fingertip recordings at 30 Hz under sedentary laboratory conditions; and MIMIC-III Waveform Database [31], containing high-resolution clinical PPG from ICU monitors at 125 Hz.

Participants were stratified into three age groups: Young (Y, 0–39 years), Middle-aged (M, 40–59 years), and Elderly (E, 60–91 years). Table 1 summarizes the demographic distribution across datasets. The three datasets exhibit markedly different age compositions: DaLiA contains no elderly participants, BUT-PPG is heavily skewed toward younger adults (71.7%), and MIMIC provides the most balanced representation across all three age strata. Age boundaries were selected to align with commonly used geriatric thresholds in clinical research and to ensure sufficient sample sizes within each stratum for stable subgroup evaluation across transfer scenarios.

**Table 1.**
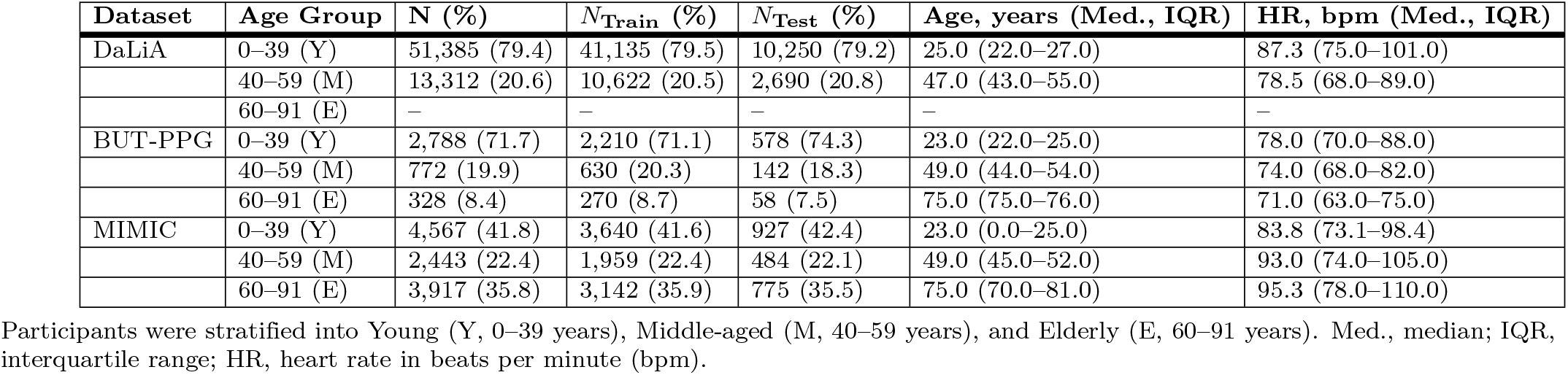
Demographic characteristics and heart rate distributions by age group across the three PPG datasets.

| Dataset | Age Group | N (%) | $N_{\text{Train}}$ (%) | $N_{\text{Test}}$ (%) | Age, years (Med., IQR) | HR, bpm (Med., IQR) |
| --- | --- | --- | --- | --- | --- | --- |
| DaLiA | 0–39 (Y) | 51,385 (79.4) | 41,135 (79.5) | 10,250 (79.2) | 25.0 (22.0–27.0) | 87.3 (75.0–101.0) |
|  | 40–59 (M) | 13,312 (20.6) | 10,622 (20.5) | 2,690 (20.8) | 47.0 (43.0–55.0) | 78.5 (68.0–89.0) |
|  | 60–91 (E) | – | – | – | – | – |
| BUT-PPG | 0–39 (Y) | 2,788 (71.7) | 2,210 (71.1) | 578 (74.3) | 23.0 (22.0–25.0) | 78.0 (70.0–88.0) |
|  | 40–59 (M) | 772 (19.9) | 630 (20.3) | 142 (18.3) | 49.0 (44.0–54.0) | 74.0 (68.0–82.0) |
|  | 60–91 (E) | 328 (8.4) | 270 (8.7) | 58 (7.5) | 75.0 (75.0–76.0) | 71.0 (63.0–75.0) |
| MIMIC | 0–39 (Y) | 4,567 (41.8) | 3,640 (41.6) | 927 (42.4) | 23.0 (0.0–25.0) | 83.8 (73.1–98.4) |
|  | 40–59 (M) | 2,443 (22.4) | 1,959 (22.4) | 484 (22.1) | 49.0 (45.0–52.0) | 93.0 (74.0–105.0) |
|  | 60–91 (E) | 3,917 (35.8) | 3,142 (35.9) | 775 (35.5) | 75.0 (70.0–81.0) | 95.3 (78.0–110.0) |

Participants were stratified into Young (Y, 0–39 years), Middle-aged (M, 40–59 years), and Elderly (E, 60–91 years). Med., median; IQR, interquartile range; HR, heart rate in beats per minute (bpm).

Heart rate distributions differ substantially both across datasets and across age groups within each dataset. In MIMIC, median heart rate increases progressively with age (Y: 83.8 bpm, M: 93.0 bpm, E: 95.3 bpm), accompanied by widening IQRs that reflect hemodynamic instability in critically ill older patients. BUT-PPG exhibits the opposite pattern: elderly participants show the lowest median heart rate (71.0 bpm) and the narrowest IQR (63–75 bpm), while younger participants display higher and more variable rates (78.0 bpm, IQR 70–88). DaLiA, recorded during physical activities, produces the highest overall heart rates (Y: 87.3 bpm, IQR 75–101) and the widest variability range across both age groups, consistent with exercise-driven cardiovascular demand.

### Bias Mitigation Strategies

We evaluated three fairness interventions during fine-tuning, each targeting a different mechanism of demographic bias correction. Inverse-frequency weighting (IF) assigns sampling weights inversely proportional to age-group frequency (Kamiran and Calders) [32]. Group distributionally robust optimization (GroupDRO) dynamically upweights the worst-performing age group during training (Sagawa et al.) [9].

Adversarial debiasing (ADV) trains a gradient-reversal adversary to prevent age-group information from encoding in learned representations (Zhang et al.) [10]. All strategies were compared against unbalanced fine-tuning as the baseline. Full mathematical formulations are provided in Appendix S1 Appendix.

### Cross-Domain Evaluation Design

To assess whether fairness interventions generalize across clinical contexts — the central question of this study — we evaluated all pairwise dataset combinations, yielding nine transfer scenarios organized into four categories:

- **Intra-domain** adaptation isolates the effect of fairness interventions from domain shift by training and testing on the same dataset with an 80/20 split.
- **Cross-domain** adaptation:
  – **Non-ICU to Non-ICU** transfer evaluates generalization between consumer and laboratory contexts, where both source and target represent healthy populations but differ in device type, recording protocol, and activity level.
  – **Non-ICU to ICU** transfer tests whether models fine-tuned on healthy populations retain clinical utility when deployed in intensive care, introducing simultaneous shifts in device, population health status, and age-performance relationships.
  – **ICU to non-ICU** transfer evaluates whether clinical optimization creates transferable representations or embeds pathology-specific features that interfere with wellness monitoring.

This design enables systematic assessment of how fairness interventions interact with progressively larger distribution shifts, from matched contexts through cross-device transfer to cross-pathology deployment.

### Evaluation Metrics

We report mean absolute error (MAE) in beats per minute for overall and age-stratified prediction performance. The fairness gap (FG) is defined as the mean of absolute pairwise MAE differences across age groups.

To summarize intervention effects systematically, we classified each method–context combination by the joint direction of MAE and FG change relative to the unbalanced baseline into one of four outcome types: Genuine improvement (G), where both MAE and FG decreased; Leveling down (L), where FG decreased but MAE increased — indicating disparity reduction through accuracy degradation of lower-error groups; Selective benefit (S), where MAE decreased but FG increased — indicating accuracy gains concentrated in already lower-error groups; and Both worse (W), where both MAE and FG increased.

To assess whether fairness interventions alter the relative difficulty structure across age groups, we compare age-group MAE rankings (Y, M, E) before and after each intervention within each transfer scenario. Cases where the intervention changes which age group exhibits the highest or lowest prediction error are reported as ordering reversals, indicating structural distortion beyond aggregate FG changes.

To characterize how fairness interventions alter learned representations, we computed silhouette scores measuring age-group clustering coherence and average inter-group Euclidean distances between age-group centroids, both computed on PCA-reduced embeddings (50 components) extracted from the model’s penultimate layer using the full test cohort (Tables 2–5); PCA was selected over nonlinear projections because it preserves global distance structure required for silhouette and inter-group distance computation [33]. For visualization, embeddings were projected to two dimensions via PaCMAP [34], which preserves local neighborhood structure but does not preserve inter-cluster distances and is therefore unsuitable for quantitative metrics; PaCMAP projections using gender-balanced subsamples filtered to overlapping heart rate ranges are provided for qualitative inspection only (Appendix Figures S1 Appendix.

**Table 2.** Intra-domain performance and fairness metrics across fine-tuning strategies.

| Dataset | Scheme | MAE |  |  |  | Fairness & Embedding |  |  | Type |
| --- | --- | --- | --- | --- | --- | --- | --- | --- | --- |
|  |  | Overall | Y | M | E | FG | Silhou. | Inter-Dist |  |
| DaLiA | Unbal. | **3.87** | **3.96** | **3.52** | – | 0.44 | −0.057 | 111.69 | – |
|  | IF | 4.34↑ | 4.62 | 3.32 | – | 1.30↑ | −0.051 | 104.73 | W |
|  | DRO | 5.28↑ | 5.27 | 5.30 | – | **0.029**↓ | −0.065 | 104.04 | L |
|  | ADV | 4.86↑ | 4.87 | 4.86 | – | **0.0099**↓ | −0.062 | 14.08 | L |
| BUT-PPG | Unbal. | **10.04** | **10.80** | **8.20** | 7.12 | 2.45 | 0.032 | 75.08 | – |
|  | IF | 10.17↑ | 11.19 | 8.23 | **4.81** | 4.25↑ | 0.038 | 65.26 | W |
|  | DRO | 10.69↑ | 10.88 | 9.83 | 11.03 | **0.80**↓ | 0.024 | 79.35 | L |
|  | ADV | 10.18↑ | 10.80 | 8.54 | 8.09 | 1.80↓ | 0.037 | 21.66 | L |
| MIMIC | Unbal. | 10.11 | 9.41 | 8.44 | 11.98 | 2.36 | −0.002 | 2.07 | – |
|  | IF | **9.88**↓ | 9.63 | **7.25** | **11.84** | 3.06↑ | −0.009 | 2.61 | S |
|  | DRO | 11.48↑ | 11.25 | 10.21 | 12.56 | **1.57**↓ | −0.001 | 1.81 | L |
|  | ADV | 9.90↓ | **9.08** | 8.18 | 11.97 | 2.50↑ | −0.013 | 0.38 | S |

Y, Young (0–39 years); M, Middle-aged (40–59 years); E, Elderly (60–91 years). Silhou., silhouette score; Inter-Dist, average inter-group Euclidean distance between age-group centroids. IF, inverse-frequency weighting; DRO, group distributionally robust optimization; ADV, adversarial debiasing. Bold values indicate the best (lowest) result per metric within each dataset; dashes indicate absent age groups. FG, fairness gap (mean absolute pairwise MAE difference across age groups). Arrows (↑/↓) on Overall MAE and FG indicate change relative to unbalanced baseline: ↓ = improved, ↑ = worsened. Type column: W = both metrics worsened; L = fairness improved but accuracy worsened (leveling); S = accuracy improved but fairness worsened (selective).

## Results

### Baseline Age-Group Performance Rankings Vary Across Clinical Contexts

Before evaluating fairness interventions, we characterized baseline age-group performance under standard (unbalanced) fine-tuning across three distinct clinical contexts (Table 2). The relative performance across age groups — which group exhibits highest versus lowest prediction error — was not consistent across datasets but instead reversed depending on the physiological context.

In the consumer wearable setting (DaLiA), which contained only Young and Middle-aged participants, the minority Middle-aged group (20.6%) outperformed the majority Young group (MAE: 3.52 vs. 3.96 bpm), consistent with lower heart rate variability during daily activities (IQR: 68–89 vs. 75–101 bpm). In the laboratory-controlled setting (BUT-PPG), the Elderly group achieved the lowest MAE (7.12 bpm) despite comprising only 8.4% of the training data, while the majority Young group (71.7%) exhibited the highest error (10.80 bpm). This pattern reflected protocol-specific physiology: sedentary recording conditions under which elderly participants demonstrated the narrowest heart rate range (IQR: 63–75 bpm), creating the simplest prediction target despite minimal sample representation. In the ICU setting (MIMIC), this hierarchy reversed: the Elderly group — the second-largest cohort (35.8%) — exhibited the highest MAE (11.98 bpm), while Middle-aged patients (22.4%) achieved the lowest error (8.44 bpm). Unlike the non-ICU settings, this ranking was not aligned with sample size. Instead, it reflected the severity of age-dependent hemodynamic instability in critically ill patients: Elderly patients exhibited the widest heart rate range (IQR: 78–110 bpm) and the highest median HR (95.3 bpm), consistent with exhausted autonomic reserves and erratic hemodynamic fluctuation in this population.

Across all three contexts, the relative prediction difficulty across age groups aligned with context-specific physiological structure rather than with demographic representation or sample size — the same group (Elderly) that was easiest to predict in a sedentary laboratory setting became the hardest in the ICU. These inversions indicate that age-based performance disparities reflect context-dependent physiological heterogeneity rather than a stable, correctable bias.

### No Intervention Simultaneously Improved Both Accuracy and Fairness

We then examined how fairness interventions (IF, GroupDRO, ADV) altered the age-group performance established in Section and shown in Table 2. Across all nine method–context combinations, no intervention simultaneously reduced overall MAE and FG relative to unbalanced fine-tuning. Instead, every combination fell into one of three outcome types: both accuracy and fairness worsened (W, 2 of 9), accuracy worsened while fairness improved — a pattern consistent with leveling (L, 5 of 9) — or accuracy improved while fairness worsened, indicating selective benefit to already-lower-error groups (S, 2 of 9).

#### FG reduction was the most common outcome but was always accompanied by overall accuracy degradation

In five of nine combinations, interventions reduced FG while increasing overall MAE. GroupDRO exhibited this pattern across all three contexts (DaLiA: MAE 3.87 to 5.28, FG 0.44 to 0.029; BUT-PPG: MAE 10.04 to 10.69, FG 2.45 to 0.80; MIMIC: MAE 10.11 to 11.48, FG 2.36 to 1.57), and ADV did so in DaLiA (MAE 3.87 to 4.86, FG 0.44 to 0.0099) and BUT-PPG (MAE 10.04 to 10.18, FG 2.45 to 1.80).

In each case, per-group analysis revealed that FG reduction was achieved by raising errors for lower-error groups rather than lowering errors for higher-error groups. In BUT-PPG under GroupDRO, this effect was most extreme: Elderly MAE increased from 7.12 to 11.03 bpm, shifting Elderly from the lowest-error to the highest-error group and reversing the baseline ranking entirely (E < M < Y became M < Y < E). In DaLiA, GroupDRO and ADV compressed both groups to near-identical error levels (DRO: Young 5.27, Middle-aged 5.30; ADV: Young 4.87, Middle-aged 4.86 bpm), effectively eliminating performance differences at the cost of 36–26% MAE increases. In MIMIC, GroupDRO preserved the baseline ranking (M < Y < E) but worsened all groups, with the largest absolute increases for Young (+1.84 bpm) and Middle-aged (+1.77 bpm).

#### IF worsened both accuracy and fairness in two of three contexts

In DaLiA and BUT-PPG, IF increased overall MAE while simultaneously widening FG (DaLiA: MAE 3.87 to 4.34, FG 0.44 to 1.30; BUT-PPG: MAE 10.04 to 10.17, FG 2.45 to 4.25). In both cases, IF preferentially reduced error for the already-lowest-error group: Middle-aged in DaLiA (3.52 to 3.32 bpm) and Elderly in BUT-PPG (7.12 to 4.81 bpm), while worsening the highest-error group. The BUT-PPG case produced the largest intra-domain FG observed across all settings (4.25 bpm).

#### Overall accuracy improvement occurred in two cases but without fairness benefit

IF in MIMIC (MAE 10.11 to 9.88) and ADV in MIMIC (MAE 10.11 to 9.90) both improved overall accuracy, but FG widened in each case (IF: 2.36 to 3.06; ADV: 2.36 to 2.50). Both interventions preferentially improved lower-error groups — IF reduced Middle-aged error from 8.44 to 7.25 bpm, and ADV improved Young (9.41 to 9.08) and Middle-aged (8.44 to 8.18) — while Elderly error remained effectively unchanged (11.98 to 11.84 and 11.97, respectively). These accuracy gains thus selectively benefited groups that already performed better, widening the gap to the highest-error Elderly group.

#### Embedding changes did not correspond to fairness outcomes

Across all method–context combinations, changes in embedding structure did not reliably predict fairness improvement or degradation (Table 2 and Appendix Figure S1 Appendix). In BUT-PPG, positive silhouette scores (0.024 to 0.038) persisted across all methods, yet fairness outcomes ranged from the worst observed (IF: FG = 4.25) to the best (DRO: FG = 0.80). In MIMIC, all methods produced near-zero silhouette scores (*−*0.001 to*−*0.013) indicating overlapping age-group clusters, regardless of whether FG improved or worsened.

These results indicate that fairness interventions, when applied to age-based performance disparities rooted in context-dependent physiology, consistently produced tradeoffs between accuracy and fairness rather than joint improvements. FG reduction was achievable but occurred through leveling — degrading performance for lower-error groups — rather than through improving predictions for higher-error groups.

### Cross-Domain Transfer Partially Restores Accuracy–Fairness Tradeoffs

We next assessed whether the accuracy–fairness tradeoffs observed within domains (Section) persisted under cross-domain transfer (Tables 3–5). Across eighteen cross-domain method–context combinations, the dominant outcome shifted: both accuracy and fairness worsened simultaneously in nine of eighteen cases, fairness improved at the cost of accuracy in five (leveling), and four achieved genuine simultaneous improvement in both MAE and FG. This contrasts sharply with intra-domain results, where no combination achieved genuine improvement (0 of 9). However, the distribution of outcomes depended heavily on transfer direction and source context.

**Table 3.** Cross-domain non-ICU transfer: age-stratified MAE (bpm), FG, and embedding metrics between BUT-PPG and DaLiA.

| Dataset | Scheme | MAE |  |  |  | Fairness & Embedding |  |  | Type |
| --- | --- | --- | --- | --- | --- | --- | --- | --- | --- |
|  |  | Overall | Y | M | E | FG | Silhou. | Inter-Dist |  |
| *BUT-PPG* → *DaLiA* | $N_{\text{train}}$ (%) | | 2,788 (71.7) | 772 (19.9) | 328 (8.4) | | | | |
| | $N_{\text{test}}$ (%) | | 51,385 (79.4) | 13,312 (20.6) | – | | | | |
|  | Unbal. | 18.81 | 20.23 | **13.33** | – | 6.90 | −0.017 | 2.57 | – |
|  | IF | 18.49↓ | 19.72 | 13.79 | – | 5.92↓ | −0.011 | 3.35 | G |
|  | DRO | **17.74**↓ | **18.28** | 15.71 | – | **2.56**↓ | −0.015 | 3.83 | G |
|  | ADV | 19.03↑ | 20.28 | 14.24 | – | 6.03↓ | −0.009 | 1.59 | L |
| *DaLiA* → *BUT-PPG* | $N_{\text{train}}$ (%) | | 51,385 (79.4) | 13,312 (20.6) | – | | | | |
| | $N_{\text{test}}$ (%) | | 2,788 (71.7) | 772 (19.9) | 328 (8.4) | | | | |
|  | Unbal. | **10.68** | **11.63** | **8.59** | **7.49** | **2.70** | 0.061 | 40.62 | – |
|  | IF | 13.16↑ | 14.04 | 11.49 | 9.67 | 2.91↑ | −0.006 | 58.89 | W |
|  | DRO | 14.67↑ | 15.68 | 13.21 | 9.62 | 4.04↑ | 0.092 | 31.20 | W |
|  | ADV | 14.62↑ | 15.36 | 13.45 | 11.11 | 2.83↑ | 0.082 | 5.58 | W |

#### In non-ICU transfer, DaLiA as source produced uniformly negative outcomes, while BUT-PPG as source permitted genuine improvement (Table 3)

When transferring from DaLiA to BUT-PPG, all three interventions worsened both MAE and FG relative to unbalanced fine-tuning: IF (MAE 10.68 to 13.16, FG 2.70 to 2.91), GroupDRO (MAE 10.68 to 14.67, FG 2.70 to 4.04), and ADV (MAE 10.68 to 14.62, FG 2.70 to 2.83). The unbalanced model also generalized to BUT-PPG’s Elderly group (MAE: 7.49 bpm), a population entirely absent from DaLiA training data, outperforming balanced methods’ predictions for groups that were explicitly represented during training. In the reverse direction (BUT-PPG *→*DaLiA), two of three interventions achieved genuine improvement: IF reduced both MAE (18.81 to 18.49) and FG (6.90 to 5.92), and GroupDRO achieved more substantial improvement (MAE 18.81 to 17.74, FG 6.90 to 2.56). ADV improved fairness (6.90 to 6.03) but slightly worsened accuracy (18.81 to 19.03). All methods nonetheless exhibited high absolute error (17.74–19.03 bpm), reflecting a 4.6–4.9 *×* increase over intra-domain DaLiA performance.

#### In non-ICU to ICU transfer, GroupDRO achieved genuine improvement in both source directions (Table 4)

**Table 4.** Cross-domain non-ICU to ICU transfer: age-stratified MAE (bpm), FG, and embedding metrics for BUT-PPG and DaLiA fine-tuned models evaluated on MIMIC.

| Dataset | Scheme | MAE |  |  |  | Fairness & Embedding |  |  | Type |
| --- | --- | --- | --- | --- | --- | --- | --- | --- | --- |
|  |  | Overall | Y | M | E | FG | Silhou. | Inter-Dist |  |
| *BUT-PPG → MIMIC* | $N_{\text{train}}$ (%) | | 2,788 (71.7) | 772 (19.9) | 328 (8.4) | | | | |
| | $N_{\text{test}}$ (%) | | 4,567 (41.8) | 2,443 (22.4) | 3,917 (35.8) | | | | |
|  | Unbal. | 25.12 | 21.19 | 25.54 | 29.46 | 5.59 | −0.073 | 3.12 | – |
| | IF | 26.91 ↑ | 23.22 | 26.47 | 31.50 | 5.52 ↓ | −0.057 | 3.82 | L |
|  | DRO | **23.28 ↓** | **19.44** | **24.14** | **27.22** | **5.18 ↓** | −0.062 | 3.01 | G |
| | ADV | 25.85 ↑ | 21.89 | 26.32 | 30.19 | 5.53 ↓ | −0.085 | 9.92 | L |
| *DaLiA → MIMIC* | $N_{\text{train}}$ (%) | | 51,385 (79.4) | 13,312 (20.6) | – | | | | |
| | $N_{\text{test}}$ (%) | | 4,567 (41.8) | 2,443 (22.4) | 3,917 (35.8) | | | | |
|  | Unbal. | 23.45 | 19.37 | 25.28 | 27.09 | 5.14 | −0.033 | 4.09 | – |
| | IF | 23.72 ↑ | 19.78 | 24.60 | 27.79 | 5.33 ↑ | −0.041 | 8.69 | W |
|  | DRO | **22.12 ↓** | **18.62** | **22.43** | **26.02** | **4.93 ↓** | −0.032 | 3.83 | G |
| | ADV | 25.85 ↑ | 21.97 | 25.97 | 30.31 | 5.56 ↑ | −0.042 | 1.03 | W |

Both non-ICU sources produced substantial degradation when deployed to ICU (overall MAE: 22–27 bpm vs. 10.11 bpm intra-domain MIMIC). GroupDRO achieved genuine improvement from both DaLiA (MAE 23.45 to 22.12, FG 5.14 to 4.93) and BUT-PPG (MAE 25.12 to 23.28, FG 5.59 to 5.18). IF and ADV from DaLiA worsened both metrics (IF: MAE 23.45 to 23.72, FG 5.14 to 5.33; ADV: MAE 23.45 to 25.85, FG 5.14 to 5.56), while IF and ADV from BUT-PPG worsened accuracy but marginally improved FG (IF: FG 5.59 to 5.52; ADV: FG 5.59 to 5.53). Notably, BUT-PPG *→*MIMIC transfer produced worse baseline performance than DaLiA *→*MIMIC (unbalanced MAE: 25.12 vs. 23.45 bpm) despite BUT-PPG containing Elderly participants (8.4%) absent from DaLiA. This is consistent with the hierarchy inversion established in Section : models that learned Elderly-as-lowest-error (BUT-PPG’s ranking) transferred worse to a context where Elderly-as-highest-error (MIMIC’s ranking) than models that learned no Elderly-specific features at all.

#### In ICU to non-ICU transfer, all interventions worsened accuracy; fairness gains occurred only through catastrophic degradation (Table 5)

**Table 5.** Cross-domain ICU to non-ICU transfer: age-stratified MAE (bpm), FG, and embedding metrics for MIMIC fine-tuned models evaluated on BUT-PPG and DaLiA.

| Dataset | Scheme | MAE |  |  |  | Fairness & Embedding |  |  | Type |
| --- | --- | --- | --- | --- | --- | --- | --- | --- | --- |
|  |  | Overall | Y | M | E | FG | Silhou. | Inter-Dist |  |
| *MIMIC* → *BUT-PPG* | $N_{\text{train}}$ (%) | | 4,567 (41.8) | 2,443 (22.4) | 3,917 (35.8) | | | | |
| | $N_{\text{test}}$ (%) | | 2,788 (71.7) | 772 (19.9) | 328 (8.4) | | | | |
|  | Unbal. | **14.61** | **14.19** | **13.74** | **20.27** | 4.34 | 0.053 | 22.52 | – |
|  | IF | 23.23 ↑ | 23.39 | 20.54 | 28.26 | 5.14 ↑ | 0.061 | 22.26 | W |
|  | DRO | 26.31 ↑ | 26.68 | 25.39 | 25.43 | 0.85 ↓ | 0.052 | 23.49 | L |
|  | ADV | 36.72 ↑ | 36.62 | 36.76 | 37.56 | **0.63** ↓ | 0.067 | 3.79 | L |
| *MIMIC* → *DaLiA* | $N_{\text{train}}$ (%) | | 4,567 (41.8) | 2,443 (22.4) | 3,917 (35.8) | | | | |
| | $N_{\text{test}}$ (%) | | 51,385 (79.4) | 13,312 (20.6) | – | | | | |
|  | Unbal. | **17.60** | **18.00** | **16.08** | – | **1.91** | –0.003 | 7.14 | – |
|  | IF | 23.74 ↑ | 25.30 | 17.75 | – | 7.55 ↑ | –0.004 | 8.47 | W |
|  | DRO | 28.35 ↑ | 30.11 | 21.60 | – | 8.50 ↑ | –0.004 | 7.30 | W |
|  | ADV | 41.38 ↑ | 43.37 | 33.72 | – | 9.64 ↑ | –0.004 | 1.39 | W |

MIMIC-sourced models exhibited moderate baseline degradation when deployed to healthy populations (MAE: 14.61–17.60 bpm), and all six MIMIC-source combinations produced worse overall MAE than unbalanced fine-tuning. In MIMIC *→*DaLiA, both accuracy and fairness worsened for all three interventions: IF increased MAE by 35% (17.60 to 23.74) with FG rising from 1.91 to 7.55, GroupDRO by 61% (to 28.35, FG 8.50), and ADV by 135% (to 41.38, FG 9.64). In MIMIC*→* BUT-PPG, GroupDRO and ADV achieved low FG (0.85 and 0.63 bpm) but at costs of 11.70 and 22.11 bpm additional MAE respectively — reducing disparity by uniformly degrading all groups. IF worsened both metrics (MAE 14.61 to 23.23, FG 4.34 to 5.14). These results indicate that ICU-trained age associations become uniformly harmful when deployed to healthy populations where age carries different physiological significance.

In summary across transfer scenarios, the four genuine improvements — where interventions simultaneously reduced MAE and FG — shared two features: none used MIMIC as a source, and three of four involved GroupDRO. By contrast, all six MIMIC-source transfers produced worse accuracy regardless of intervention, and the two that achieved FG improvement did so through catastrophic performance collapse (MAE exceeding 25 bpm). These directional asymmetries suggest that cross-domain fairness outcomes depend less on the intervention itself than on the alignment between source-trained performance rankings and target-domain physiological structure.

## Discussion

Our results indicate that age-group performance hierarchies in PPG-based heart rate prediction are context-dependent and resist correction through standard fairness interventions, with gap reduction occurring predominantly through accuracy degradation rather than genuine subgroup improvement.

### Fairness gap reduction was achieved predominantly through leveling down rather than genuine improvement

In intra-domain settings, five of nine method–context combinations reduced FG while simultaneously increasing overall MAE — a pattern consistent with leveling down as formalized by Mittelstadt et al. [14]. This proportion parallels findings in chest X-ray classification, where Zhang et al. [15] reported that all nine fairness methods achieving group fairness did so by worsening performance across all groups, and Pfohl et al. [16] characterized the phenomenon as “nearly-universal degradation.”

Our results extend the leveling-down observation to a regression setting (heart rate prediction from PPG signals) and to the age attribute, which differs structurally from the race and sex attributes typically studied. The 2*×* 2 outcome classification makes the mechanism explicit: reduced FG co-occurring with increased MAE indicates that the disparity metric improved not because disadvantaged groups benefited but because advantaged groups were harmed. In the two most extreme intra-domain cases — GroupDRO on DaLiA and ADV on DaLiA — interventions compressed age-group errors to near-identical levels by increasing Young MAE by 33–17% while Middle-aged MAE changed minimally. The resulting FG values (0.03 and 0.01 bpm) suggest apparent equity, but at the cost of a model that predicts worse for every subgroup. An audit that reports only FG reduction without per-group directional analysis would classify five of our nine intra-domain results as fairness improvements — obscuring that none achieved genuine benefit for any subgroup. Our results reinforce calls by Pfohl et al. [35] and Belhadj et al. [36] for decomposed subgroup reporting and floor-based fairness criteria that structurally prevent leveling down.

### Age-group difficulty hierarchies reversed across clinical contexts, indicating that performance gaps reflect physiological structure rather than correctable bias

Elderly participants exhibited the lowest MAE in BUT-PPG (laboratory setting) but the highest MAE in MIMIC (ICU) — a hierarchy inversion that, to our knowledge, has not been previously documented empirically for a physiological prediction attribute across clinical deployment contexts. This inversion indicates that age-related performance gaps reflect context-specific measurement and physiological conditions rather than a stable attribute-level bias amenable to algorithmic correction.

This distinction matters because standard fairness interventions implicitly assume that performance gaps reflect correctable artifacts — distributional imbalance, label noise, or representational shortcomings — that a reweighting or robust optimization procedure can address. When the gap instead reflects that a particular age group presents genuinely more variable physiology in a given context (e.g., elderly ICU patients with heterogeneous hemodynamic responses), interventions designed to equalize outcomes can only do so by degrading predictions for groups whose physiology is more predictable. The hierarchy inversion provides empirical evidence that the assumption of correctable bias does not hold uniformly across clinical deployment contexts, at least for the age attribute in PPG-based prediction. This observation aligns with the broader concern raised by Zhang et al. [15], who advocated for “investigation of the bias-inducing mechanisms in the underlying data distribution” before applying fairness corrections; our results suggest that such investigation should specifically examine whether performance hierarchies are stable across intended deployment contexts.

### Cross-domain transfer amplified within-domain patterns and was strongly asymmetric by source context

Intra-domain evaluation produced zero genuine improvements out of nine combinations; cross-domain evaluation produced four genuine improvements out of eighteen, but also nine combinations where both MAE and FG worsened simultaneously. All six MIMIC-source transfers worsened accuracy regardless of intervention, while all four genuine improvements originated from non-ICU source datasets. This source-context dependence is consistent with Yang et al. [17], who demonstrated that fairness corrections optimized for in-distribution data do not maintain their properties under distribution shift. Our results add a specific mechanism: when source and target contexts exhibit *inverted* performance hierarchies (e.g.,

Elderly-as-easiest in BUT-PPG vs. Elderly-as-hardest in MIMIC), fairness interventions that encode source-specific age associations transfer especially poorly. Notably, BUT-PPG*→* MIMIC transfer produced worse baseline performance than DaLiA*→* MIMIC despite BUT-PPG containing Elderly participants that DaLiA lacked, suggesting that learning inverted age–performance associations is more harmful than learning no age-specific features at all.

The four genuine improvements all involved GroupDRO (three of four) and non-MIMIC sources. GroupDRO’s minimax formulation — optimizing worst-group performance rather than equalizing across groups — may be less susceptible to encoding context-specific hierarchies than methods that explicitly target inter-group equalization. However, GroupDRO also produced leveling-down outcomes in all three intra-domain contexts, indicating this advantage is transfer-specific rather than a general property of the method.

### Representational separation did not predict fairness outcomes

Embedding analysis (Appendix S1 Appendix) indicated that representational geometry did not predict fairness outcomes. ADV systematically compressed inter-group distances (lowest in 8 of 9 scenarios), consistent with its adversarial objective, but this compression did not translate to fairness improvement — ADV produced leveling-down outcomes in both non-ICU intra-domain settings and achieved its lowest cross-domain FG only through an MAE increase of 22.11 bpm above baseline. More broadly, similar embedding geometries coexisted with opposing fairness outcomes across datasets (e.g., in BUT-PPG intra-domain, the highest and lowest FG values occurred under high inter-group distances). These patterns suggest that age-based performance disparities are driven by the physiological data-generating process rather than by learnable representational structure amenable to intervention.

These findings carry practical implications for clinical deployment. Fairness interventions should not be assumed to transfer across clinical contexts: an intervention that reduces age-group disparities in one setting may worsen both accuracy and fairness in another when the target context exhibits different age–performance relationships.

Fairness gap reduction alone is an insufficient evaluation criterion; without per-subgroup directional analysis, apparent improvements may reflect leveling down rather than genuine equity gains. When performance hierarchies are context-dependent — as observed here across ICU, laboratory, and consumer wearable settings — maintaining domain-specific models with context-appropriate fairness evaluation may be more defensible than pursuing a single fairness-corrected model for cross-context deployment.

### Limitations

Several limitations should be considered. First, we evaluated three fairness interventions (IF, GroupDRO, ADV) across three datasets; other methods — including post-processing calibration, mixup-based augmentation, or multi-objective optimization — may behave differently. Second, our analysis is restricted to the age attribute in PPG-based heart rate prediction; the extent to which hierarchy inversion and leveling-down patterns generalize to other protected attributes or physiological signals remains an empirical question. Third, we used a single pretrained foundation model architecture, and the interaction between model capacity, pretraining strategy, and fairness intervention effects warrants further investigation. Fourth, the three datasets differ not only in clinical context but also in sample size, age distribution, and recording protocol, making it difficult to isolate which contextual factor drives the observed hierarchy inversions. Additionally, our three-group age stratification, while aligned with established geriatric thresholds, may obscure finer-grained patterns; whether hierarchy inversions persist under alternative age boundaries remains untested. Finally, our cross-domain evaluation uses held-out test sets as proxies for deployment distribution shift; real-world deployment may introduce additional sources of variation not captured by these benchmarks.

## Conclusion

This study evaluated whether fairness interventions applied during fine-tuning of a pretrained PPG model could reduce age-group performance disparities in heart rate prediction across three clinical contexts. No intervention achieved simultaneous improvement in both accuracy and fairness within any single domain (0 of 9 combinations); the dominant outcome was leveling, in which fairness gap reduction was achieved by degrading predictions for lower-error groups rather than improving them for higher-error groups (5 of 9). Age-group difficulty rankings reversed across contexts — the same demographic group shifted from lowest to highest prediction error depending on the physiological setting — suggesting that these disparities reflect context-dependent physiological structure rather than correctable algorithmic bias. Under cross-domain transfer, genuine improvement emerged in 4 of 18 combinations, exclusively from non-ICU source domains and predominantly through GroupDRO, while all ICU-source transfers worsened accuracy regardless of intervention. These findings indicate that fairness interventions designed to correct data-driven artifacts may be structurally mismatched to age-based performance disparities rooted in physiology, and that fairness evaluation in clinical ML should incorporate cross-domain assessment alongside within-domain metrics to detect leveling and context-dependent instability before deployment.

## Data Availability

The PPG-DaLiA dataset is available from the UCI Machine Learning Repository (https://archive.ics.uci.edu/dataset/495/ppg+dalia). The Brno University of Technology Smartphone PPG Database (BUT-PPG) is available from PhysioNet (https://physionet.org/content/butppg/). The MIMIC-III Waveform Database is available from PhysioNet (https://physionet.org/content/mimic3wdb/1.0/).

## Supporting information

**S1 Appendix. Supplementary methods and results**. Fine-tuning hyperparameters, mathematical formulations of bias mitigation strategies (IF, GroupDRO, ADV), and embedding structure analysis across intra-domain and cross-domain transfer scenarios.

**S1 Table. Fine-tuning hyperparameters for the 19M GPT-PPG model**. All values are shared across unbalanced and bias-aware fine-tuning strategies.

**S1 Fig. Intra-domain embedding space visualizations across fine-tuning strategies**. Each row corresponds to a dataset with train and test splits from the same source. Columns show Unbalanced, IF, DRO, and ADV methods.

**S2 Fig. Cross-domain non-ICU to non-ICU embedding space visualizations**. Top row: DaLiA-trained models evaluated on BUT-PPG. Bottom row: BUT-PPG-trained models evaluated on DaLiA.

**S3 Fig. Cross-domain non-ICU to ICU embedding space visualizations**. Top row: DaLiA-trained models evaluated on MIMIC. Bottom row: BUT-PPG-trained models evaluated on MIMIC.

**S4 Fig. Cross-domain ICU to non-ICU embedding space visualizations**. Top row: MIMIC-trained models evaluated on DaLiA. Bottom row: MIMIC-trained models evaluated on BUT-PPG.

## Acknowledgments

We acknowledge the developers of the DaLiA, BUT-PPG, and MIMIC-III waveform databases. Computational resources were provided by Emory University.

